# Characteristics of the Hypertensive Pediatric Population of Northern Israel

**DOI:** 10.1101/2025.01.03.25319982

**Authors:** Lotem Weiss, Daniella Magen, Shirley Pollack

## Abstract

**Objectives:** The overall prevalence of hypertension (HTN) among Israeli Jewish adolescents was reported to be 0.4% in males and 0.074% in females. The demographic characteristics of Northern Israeli population are unique, with a nearly equal distribution of Arab and Jewish ethnicity. We describe the demographic and clinical characteristics of pediatric hypertensive patients in northern Israel.

**Design:** Data was retrospectively collected from electronic medical records (EMR) of pediatric patients aged 0-18 diagnosed with HTN at Rambam Health Care Campus between 2010-2020. Demographic characteristics, etiology, end-organ damage (EOD), and medical treatment were collected and compared between specific subgroups.

**Results:** During the study period, 479 children diagnosed with HTN were included. Mean age at diagnosis was 9.6 years. 64% were males. BMI > 85th percentile was measured in 45%. 17% were diagnosed with primary HTN. Primary HTN was more prevalent (32%) in adolescents, the majority of Jewish ancestry. Obesity prevalence was significantly higher in primary HTN patients. EOD was more common in secondary HTN, Arab-Muslim origin, consanguinity, and younger age. At follow-up, 35.8% were without antihypertensive medications. Calcium channel blockers were the most frequently prescribed anti-hypertensives.

**Conclusions:** Higher rates of secondary HTN than reported in the literature were attributed to high consanguinity and resulted in extensive diagnostic procedures across all age groups, unlike AAP guidelines recommendations. Better adherence to guideline recommendations was demonstrated in patients referred to the pediatric nephrology institute, highlighting the importance of referral of HTN patients to specialized clinics.

## Introduction

Over the past decades, there has been a global increase in the prevalence of hypertension (HTN) in children. Pediatric HTN is defined as systolic, and or diastolic blood pressure (BP) >95^th^ percentile for age, sex, and height(1). Pediatric elevated blood pressure (EBP), previously termed “prehypertension”, is defined as systolic, and or diastolic BP in the 90-95^th^ percentile for age, sex, and height. The estimated prevalence of pediatric HTN in the US is 3-4%, with additional 9-10% of children diagnosed with EBP(1,2). High BP is more prevalent in males and in older pediatric age groups(1,3). Despite its high prevalence, pediatric HTN is often underdiagnosed(2,4). This is thought to be due to difficulty in recognition of HTN in children, given the wide variability of normal BP according to age, sex, and height percentiles. In addition, EBP in children is often dismissed as secondary to fear of medical personnel, pain, stranger anxiety, etc.(5).

HTN poses a substantial health risk, due to its association with severe end-organ damage (EOD) to kidneys, eyes, and heart. EOD risk increases with younger age at onset of HTN(6). Importantly, children with EBP and with asymptomatic HTN are also at risk of developing HTN-associated EOD(7,8). Uncontrolled HTN may cause chronic kidney disease (CKD)(9), hypertensive retinopathy, left ventricular hypertrophy (LVH) and enlargement, and reduced ejection fraction(10,11). Hypertensive children are at increased risk for HTN, metabolic syndrome and cardiovascular (CVS) disease during adulthood(7,12,13).

Pediatric HTN can be classified as primary (essential) or secondary. Childhood primary HTN is strongly correlated with overweight and obesity(14,15), with a respective two and four time risk of EBP in children with obesity (BMI 95^th^–98^th^ percentiles), and severe obesity (BMI >99^th^ percentile), compared to normal weight children(1,16,17). Rapid weight gain is also correlated with increased HTN risk(18).

Secondary HTN results from underlying medical conditions. It is more common in younger pediatric age groups, including premature infants, neonates and infants(19,20), as in other chronic diseases.

Guidelines of the American Academy of Pediatrics (AAP) for diagnosis and treatment of HTN in the pediatric population (2017)(21) recommend annual BP measurement in all children over 3 years of age, and in every medical encounter in children with risk factors for HTN, including obesity, CKD and diabetes. Children younger than 3 years of age should undergo BP monitoring in the presence of selected risk factors, including history of prematurity, congenital heart defects, primary renal disease, solid-organ transplantation or malignancies. Ambulatory blood pressure monitoring (ABPM) is recommended, especially given its better correlation with EOD(22) in the general population.

Antihypertensive medications are specifically recommended in pediatric patients with stage 2 HTN, symptomatic stage 1 HTN, stage 1 hypertension with EOD, or persistently abnormal BP after 3-6 months without pharmacological intervention(21). The most prevalent medications, used in 35% of the patients, were angiotensin-converting enzyme inhibitors/angiotensin receptor blockers (ACEi/ARBs)(2).

In contrast to the above mentioned general prevalence of pediatric HTN, its reported prevalence in Israeli Jewish adolescent males and females is only 0.4%, and 0.074%, respectively(14), with a higher prevalence of 3.5-8.3% in overweight and obese males. Of note, during recent decades there has been a respective 4^th^-fold and 20-fold increase in the reported prevalence of Israeli adolescent obesity and severe obesity(23). Moreover, a large population-based retrospective study, comparing the risk of HTN in various Israeli ethnic sub-populations, reported an increased risk of HTN in Ashkenazi-Jewish adolescent compared to their Sephardi and Oriental counterparts. However, this study was of limited applicability to the entire Israeli population, as it focused on Jewish secular adolescent Israeli Defense Force (IDF) conscripts(24).

In this study we aimed to assess the demographic and clinical characteristics of pediatric hypertensive patients within the unique population of northern Israel, who were followed and managed at the largest tertiary pediatric referral center in the north of Israel.

## Methods

We collected data from electronic medical records (EMR) of pediatric patients aged 0-18 years who were diagnosed with HTN and managed at Rambam Health Care Campus between January 1^st^ 2010 and December 31^st^ 2020.

Data collection from the patients’ files was performed using MDClone, a data extraction platform, which provides patient-level data around an index event. Follow-up data was completed manually from patients’ individual EMR.

Collected data included: (a) Demographic characteristics. (b) Medical background. (c) Diagnosis and follow-up. (d) Treatment.

Patients followed at the Pediatric Nephrology Institute (including ambulatory and hospitalized patients with hypertension, primary kidney disorders, CKD, kidney failure on dialysis therapy, and renal transplant recipients) were included in the nephrology subgroup, while patients followed at other hospital units were included in the non-nephrology subgroup.

### Definition of HTN

Before 2017, HTN was defined according to the 2004 4^th^ report on the diagnosis, evaluation, and treatment of high blood pressure in children and adolescents(26). Since 2017, HTN is defined based on the 2017 AAP guidelines(21), as systolic and/or diastolic BP at or above the 95^th^ percentile of age, sex, and height-adjusted percentiles, on three separate occasions.

### Statistical analysis

Analyses were performed in R Statistical Computing Environment, version 4.2.1 (R Core Team, 2022). Continuous data were reported as mean ± standard deviation (SD) or medians and percentiles 25 and 75 (interquartile range, IQR) when appropriate. Continuous variables are compared using t-test for two groups and ANOVA for more than two groups. Categorical variables are compared using the Kruskal-Wallis rank sum test, Pearson’s Chi-squared test, Fisher’s exact test for contingency tables (p-values were simulated using 1 million replications). The level of significance was set at p<0.05.

## Results

Between 2010-2020, 538 children aged 0-18 years of age, were diagnosed as suffering from HTN. Of the 538 patients, 59 were excluded due to lack of data, or loss to follow-up, and the remaining 479 patients were included in data analysis. Of these, 268 patients (56%) were managed at the Pediatric Nephrology institute, and 211 (44%) were followed in other hospital units, including, among others, the pediatric intensive care unit (PICU), the neonatal Intensive care unit (NICU), general pediatrics, pediatric hemato-oncology, endocrinology & diabetes, and pediatric cardiology (figure 1).

**Figure 1:**
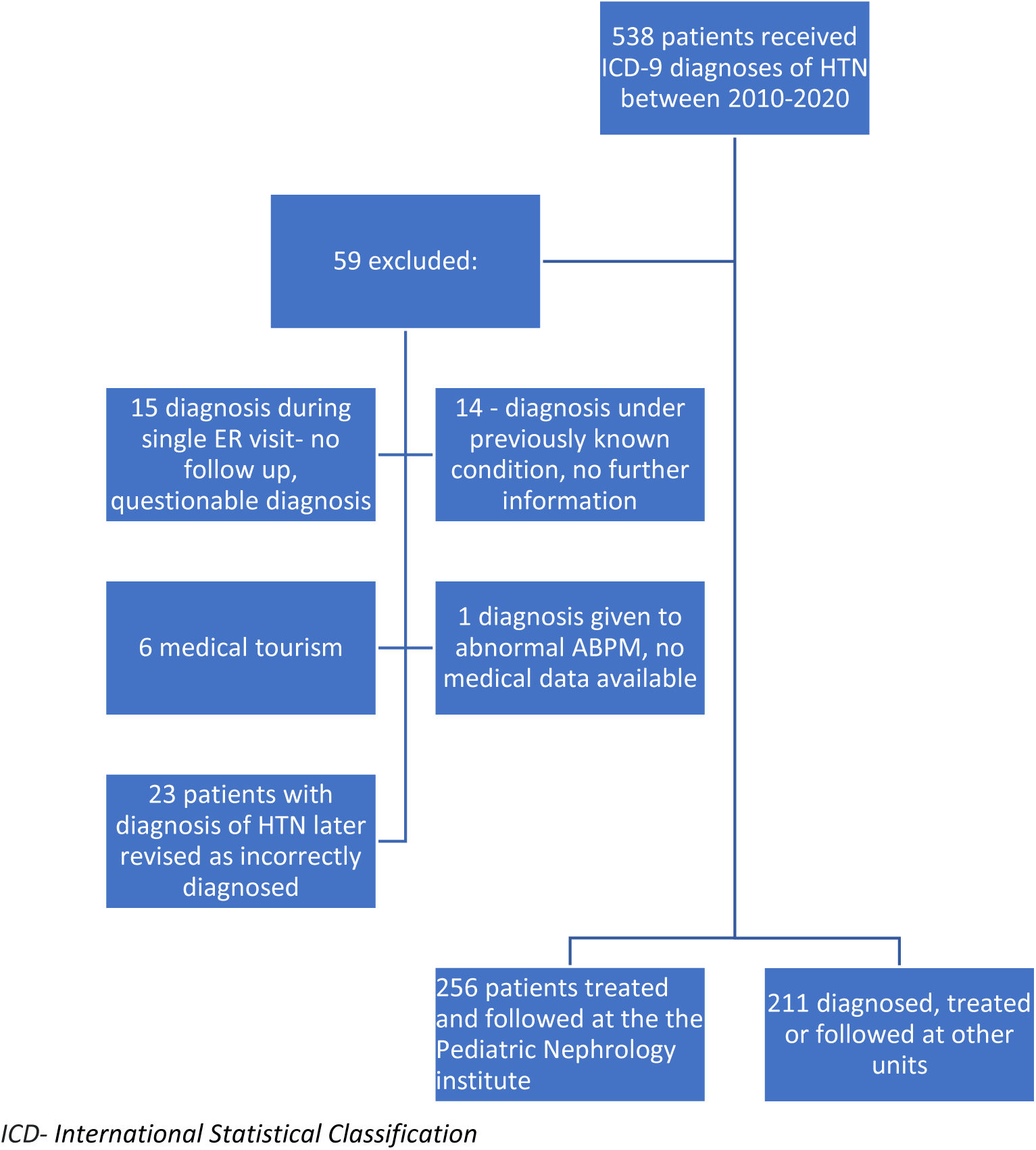
Study population.

The mean cohort age at diagnosis was 9.6 years (SD-6.3 years). There was no significant age difference between the nephrology (9.7y ± 6.1) and the non-nephrology (9.4y ± 6.6) subgroups (p=0.6). The median age at diagnosis was 11.2 years (IQR 2.8-15.6). Forty two percent of the cohort were adolescents aged 13-18 years. 64% were males, with no significant difference in gender prevalence between subgroups.

In the non-nephrology subgroup, male patients were significantly older than females (10.3 years vs. 7.9 years, respectively, p-0.017).

At the time of HTN diagnosis, 45% of the cohort had an abnormally high BMI above the 85^th^ percentile for age and sex (overweight and obesity)(Figure 2). No significant difference in BMI at diagnosis was observed between subgroups.

**Figure 2:**
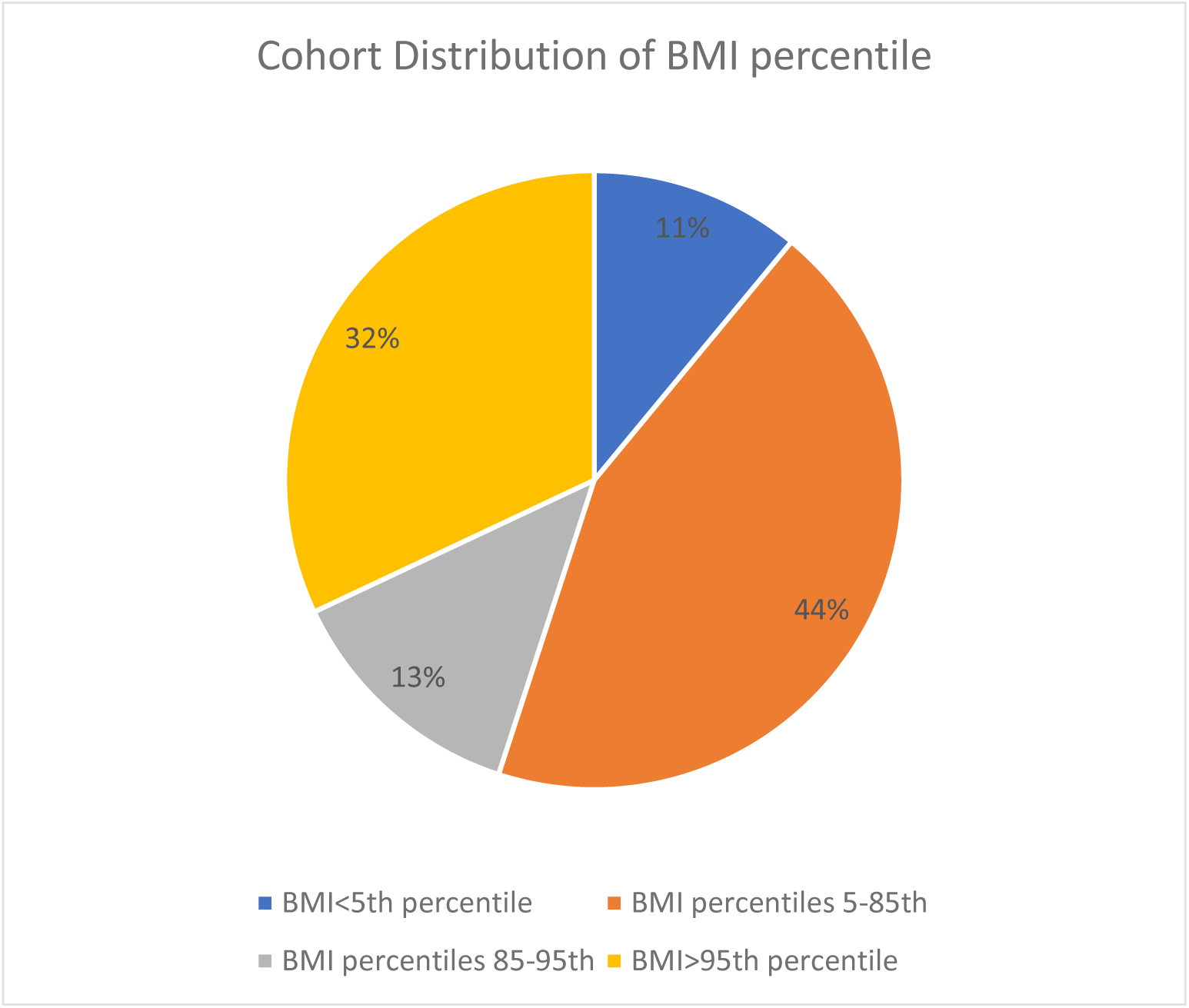
Cohort distribution of BMI percentile.

Mortality rate was significantly higher in the non-nephrology subgroup (14% vs. 6% respectively, p-0.004).

Figure 3 and 4 depicts the ethnic distribution of the entire cohort (f. 3), and of the non-nephrology subgroup (f. 4). As seen, 42% of the entire cohort, and 44% of the nephrology subgroup were of Arab-Muslim ethnicity. Thirty two percent of the entire cohort, as well as of the nephrology subgroup were composed of Jews. Arab-Christians, Druze and other ethnicities were significantly less commonly represented in all study subgroups.

**Figure 3:**
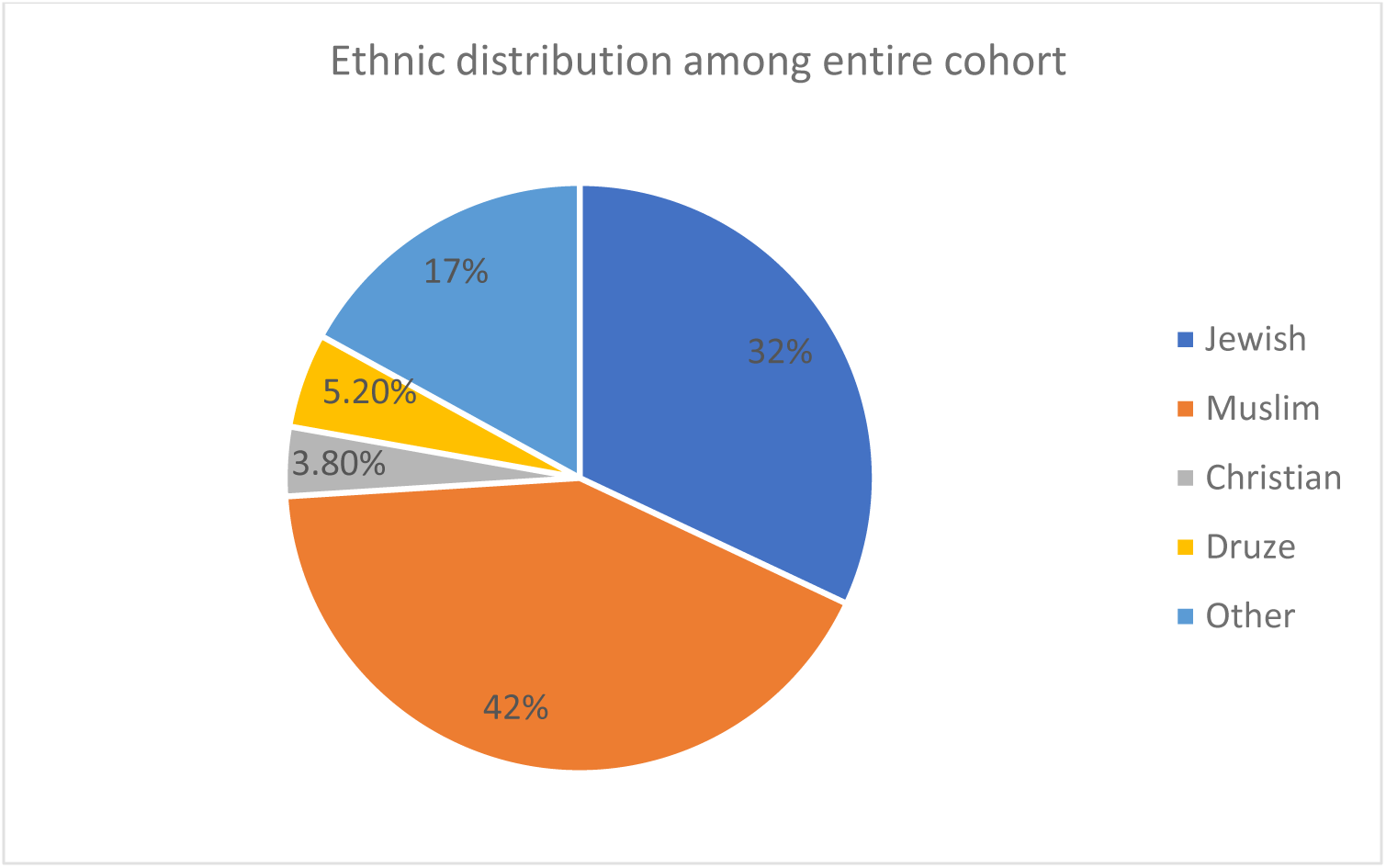
Ethnic distribution of the entire cohort.

**Figure 4:**
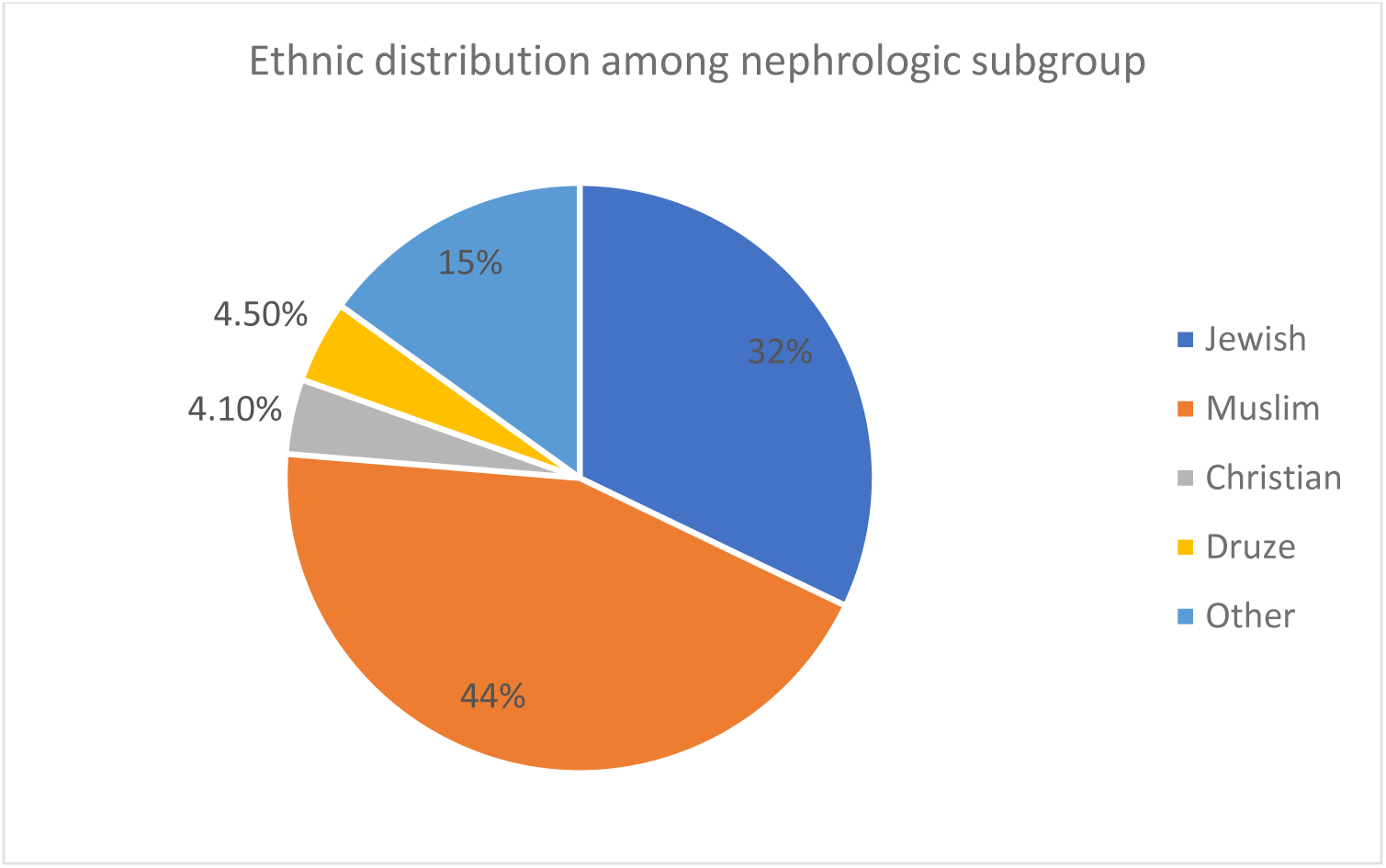
Ethnic distribution of the nephrology subgroup.

Parental consanguinity was reported in 16% of the entire cohort, with significantly higher prevalence within the nephrology compared to the non-nephrology subgroup (19% vs, 11%, respectively, p-0.022). Consanguinity was significantly more prevalent in patients of -Muslim ethnicity (30% vs. 11-16% of other ethnicities, p<0.001), in younger age groups (30% and 24% for age groups 0-1 years and 1-6 years, respectively, compared to 12% and 8.4% in older age groups, p<0.001), and in patients with secondary HTN (22% vs. 10% in patients with primary HTN, p-0.022).

Table 1 summarizes the etiologies of HTN for the entire cohort, showing significant differences in distribution of etiology between the nephrology and non-nephrology subgroups (p<0.001). Within the entire cohort, only 17% were diagnosed with primary HTN, accounting for 15% and 19% of the nephrology and non-nephrology subgroups, respectively. Overall, 62 patients (13%) were diagnosed with drug-related HTN, which was significantly more common in the non-nephrology (24%) compared to the nephrology (4.5%) subgroups. In all subgroups, the most prevalent HTN-related drugs were glucocorticoids (40 patients, 62.5% of medication-related HTN) and ACTH (9 patients, 14%), while cyclosporine A, fludrocortisone and other were less commonly reported. The main Indications for glucocorticoid administration included hemato-oncology disorders, hyper-reactive airway disease, and peri-extubation in PICU patients.

**Table 1:** Etiology distributions of HTN.

|  |  | Overall<br>N=479 | Nephrologic<br>subgroup<br>N=268 | Non-<br>nephrologic<br>subgroup<br>N=211 | p-Value<sup>1</sup> |
| --- | --- | --- | --- | --- | --- |
| Etiology | Acute N(%) | 73 (15%) | 18 (6.7%) | 55 (26%) | <0.001 |
|  | Renal N(%) | 121 (25%) | 113 (42%) | 8 (3.8%) |  |
|  | Drug-related N(%) | 63 (13%) | 12 (4.5%) | 51 (24%) |  |
|  | Vascular N(%) | 60 (13%) | 38 (14%) | 22 (10%) |  |
|  | Unknown N(%) | 39 (8.1%) | 16 (6.0%) | 23 (11%) |  |
|  | White Coat N(%) | 22 (4.6%) | 18 (6.7%) | 4 (1.9%) |  |
|  | Endocrine N(%) | 8 (1.7%) | 4 (1.5%) | 4 (1.9%) |  |
|  | Central N(%) | 3 (0.6%) | 1 (0.4%) | 2 (0.9%) |  |
|  | Primary N(%) | 80 (17%) | 40 (15%) | 40 (19%) |  |
|  | Genetic N(%) | 5 (1.0%) | 5 (1.9%) | 0 (0%) |  |
|  | Metabolic N(%) | 3 (0.6%) | 1 (0.4%) | 2 (0.9%) |  |

CAKUT= congenital anomalies of the kidney and urinary tract; ESRD = End-Stage Renal Disease
<sup>1</sup>Fisher's Exact Test for Count Data with simulated p-value (based on 1e+06 replicates); Fisher's exact test
<sup>2</sup>CoA, AS, mid aortic syndrome, RAS, NF, renal angiomyolipoma.
<sup>3</sup> Takayasu's arteritis, ANCA vasculitis, Leukocytoclastic vasculitis (Henoch-Schonlein purpura)

As expected, primary renal disorders represented the most common cause of HTN (42%) in the nephrology subgroup, with primary glomerular disease accounting for 43% of cases, followed by all-cause kidney failure in 28%, congenital anomalies of the kidneys and urinary tract (CAKUT) in 16.7%, and post renal transplantation in 12%.

Vascular etiologies for secondary HTN included aortic and main renal artery disease in 52% of patients, and vasculitis in 17%.

While prematurity accounted for 9.4% of the cohort, the prevalence of neither prematurity nor of umbilical artery catheterization differed between nephrology and non-nephrology patients (p=0.3).

EOD due to HTN (Table 2), including LVH and hypertensive retinopathy were significantly more prevalent in the nephrology compared to the non-nephrology subgroups (29% vs. 9.7% for LVH, respectively, p.<0.001; and 7.1% vs. 1.9% for hypertensive retinopathy, p<0.001). Hypertensive emergencies were also significantly more frequent in the nephrology subgroup (9.3% vs. 1.4% in non-nephrology patients, p<0.001).

**Table 2:** Complications.

| Variable | Overall<br>N=479 | Nephrologic<br>subgroup<br>N=268 | Non-<br>nephrologic<br>subgroup<br>N=211 | p-Value<sup>1</sup> |
| --- | --- | --- | --- | --- |
| Left ventricular hypertrophy (LVH) n(%) | 94 (20%) | 79 (29%) | 15 (7.1%) | <0.001 |
| Hypertensive Retinopathy n(%) | 30 (6.3%) | 26 (9.7%) | 4 (1.9%) | <0.001 |
| Dialysis during F/U n(%) | 90 (19%) | 86 (32%) | 4 (1.9%) | <0.001 |
| eGFR below 60 at F/U n(%) | 41 (22%) | 40 (28%) | 1 (2.0%) | <0.001 |
| Proteinuria (urine protein above 300) n(%) | 73 (29%) | 65 (38%) | 8 (11%) | <0.001 |
| Hypertensive emergencies n(%) | 28 (5.8%) | 25 (9.3%) | 3 (1.4%) | <0.001 |

F/U= follow-up
<sup>1</sup>Pearson's Chi-squared test

Details regarding pharmacological anti-hypertensive treatment were selectively collected for the nephrology subgroup since drug therapy documentation is inconsistent in EMR of non-nephrology patients. At the end of follow-up, 35.8% (96/268) of patients in the nephrology subgroup were not under anti-hypertensive pharmacological treatment, while 55.6% (149/268) were managed with 1-3 different anti-hypertensive medications, and 8.6% (23/268) were under 4-6 anti-hypertensive medications. Overall, 230 nephrology patients (85.8%) received at least one anti-hypertensive medication during follow-up. Of these, 79.8% (214/230) received calcium channel blockers at some point during follow-up, 35% (94/230) received β-blockers, 32.5% (87/230) received an α_2_-agonist (i.e., clonidine), 28% (75/230) received ACEi or ARBs, 13.8% (37/230) received diuretics, and 11.5% (31/230) received vasodilators, including minoxidil or hydralazine.

166 patients under the age of 6 years were included in this study. Of these, 52% (87/166) were included in the nephrology subgroup, comparable to the 56% of nephrology patients within the entire cohort. Diagnostic imaging workup, including ultrasound-Doppler, renal DMSA scan, computed tomography angiography (CTA) and/or Magnetic resonance angiography (MRA), were performed in 96 of 166 (57.8%) patients under 6 years of age in the entire cohort, and in 58 of 87 (66.66%) patients under 6 in the nephrology subgroup. Overall, imaging studies were performed in 53.4% of the entire cohort, however significantly more often (especially renal US-Doppler) in the nephrology vs. the non-nephrology subgroup (64% vs. 34%, respectively, p<0.001). ABPM was also performed significantly more often in nephrology compared to non-nephrology patients (35% vs. 5.7%, respectively, p<0.001).

During follow-up, almost 2% of the entire cohort underwent renal artery interventions, and 2.9% underwent unilateral or bilateral nephrectomy, indicated for Wilms’ tumor, or for uncontrollable drug-resistant HTN.

When comparing the characteristics of patients from different ethnic subgroups (table 3), Arab-Muslim patients were significantly younger (median age - 9 years, vs. 10.5-14.2 years in other ethnicities, p-0.018), more commonly had parental consanguinity, showed a significantly lower prevalence of primary vs. secondary HTN (11% vs. 19-33%, p-0.001), a non-significantly higher prevalence of CKD (31% with eGFR < 60 ml/min/1.73m^2^ at diagnosis, vs. 7.7-22% for other ethnicities, p-0.06), and a significantly higher complication rate, including LVH (29% vs. 11-24%, p<0.001), dialysis dependence (32% vs. 7.2-16%, p<0.001), and hyperuricemia (33% vs. 11-20%, p-0.004). Hypertensive retinopathy and hypertensive emergencies were not significantly different between ethnic groups. Of note, obesity was more prevalent in Jews and Arab-Christians compared to other ethnicities (p-0.01). Other comorbidities of HTN did not differ between ethnic groups.

**Table 3:**
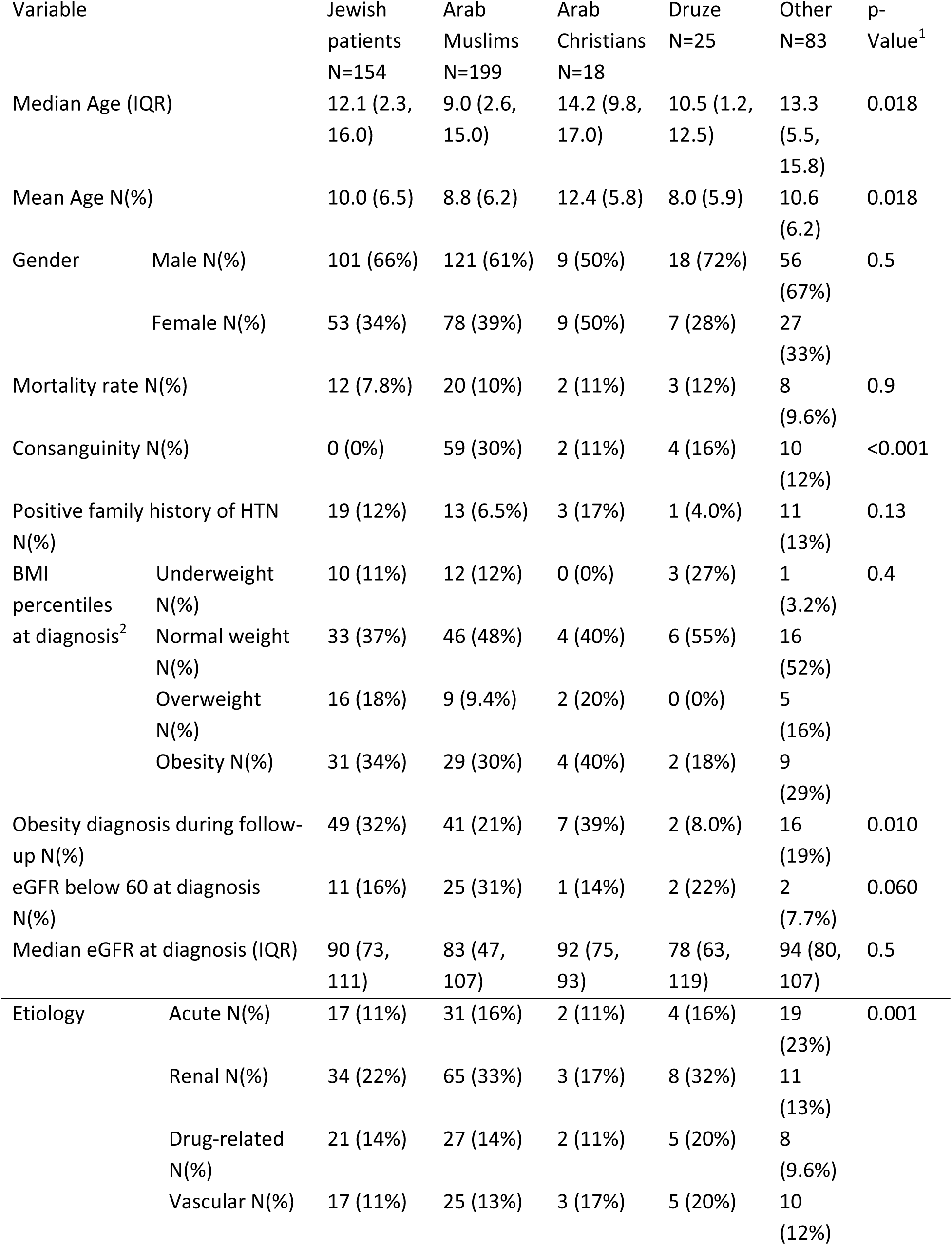
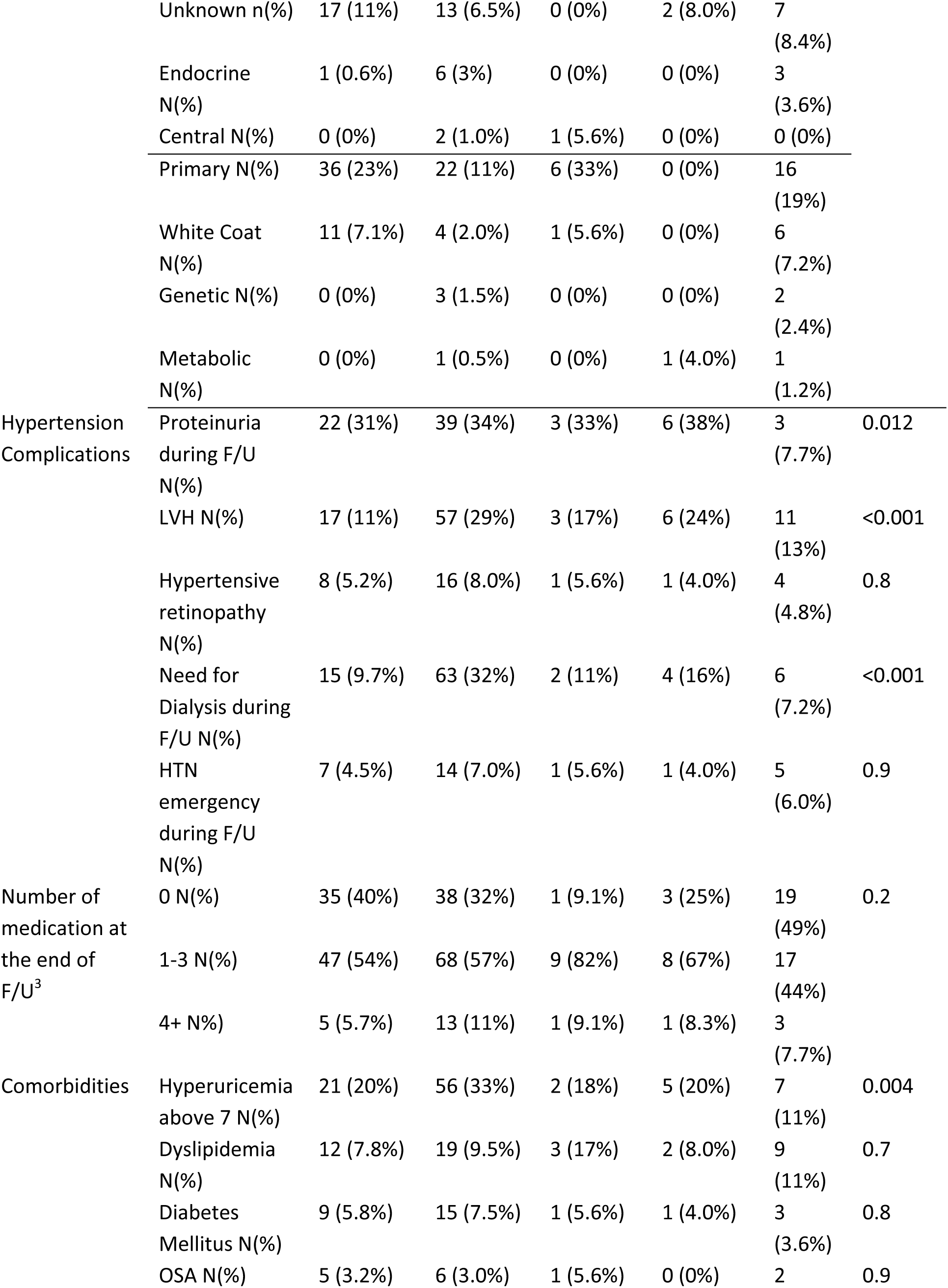
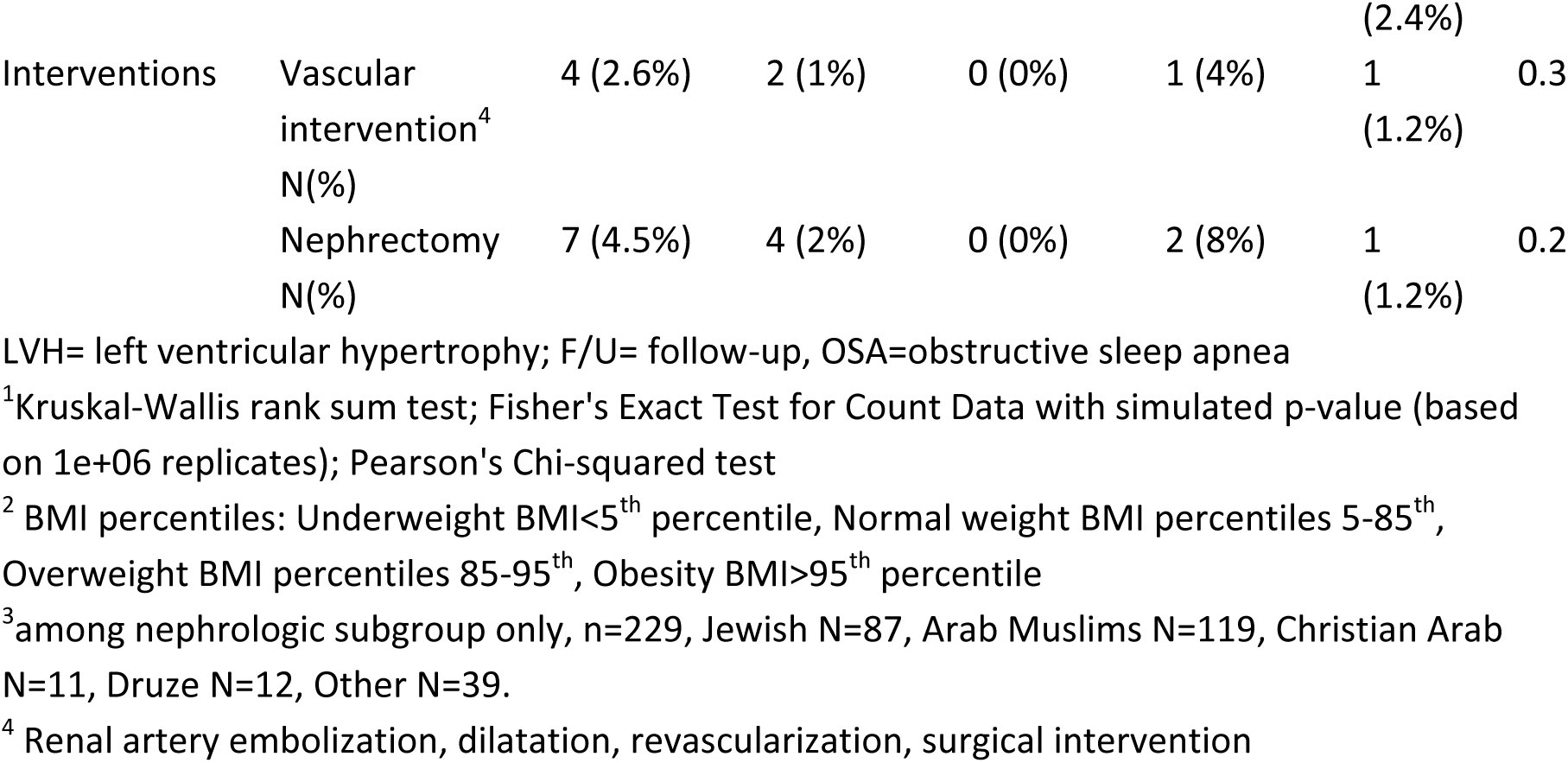
Characteristics among HTN patients of different ethnic groups.

Younger age groups at presentation of HTN were characterized by significantly higher rates of the following characteristics: (a) mortality (16% for age 0-6 years, compared to 8.2% for 6-12 years, and 4.4% for adolescents, p-0.002); (b) parental consanguinity (30% for infants aged 0-1 year, compared to 8.4% in older groups, p<0.001); (c) history of prematurity (12.6% of infants aged 0-1 years born before 30 weeks’ gestation, vs. 3% of age group 13-18 years p<0.001); (d) vascular etiology (21-22% for patients 0-6 years, vs. 6.9-10% for patients older than 6 years); (e) complications of HTN (up-to 31% LVH, compared to 14% in the 13-18 year age group, p-0.008); (f) surgical nephrectomy (6% vs. 1-2% in older age groups, p-0.008) (Figure 5).

**Figure 5:**
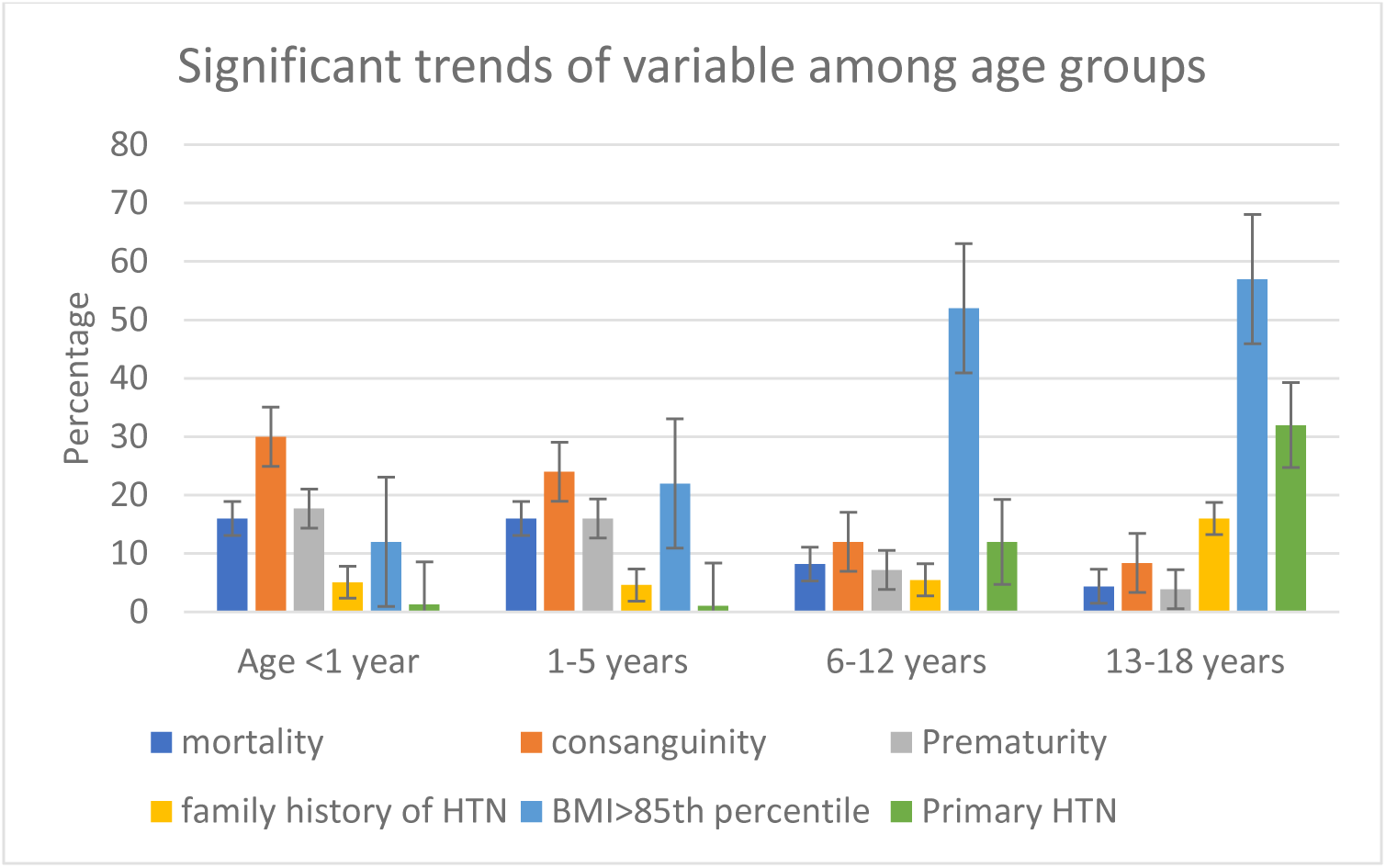
Trends of variables among different age groups.

In contrast, hypertensive adolescents aged 13-18 years showed a higher prevalence of primary HTN (32% vs. 12% for ages 6-12 years, and 1.2% under 6 years, p<0.001), and higher rates of comorbidities, including DM (11% vs. 0-4.5% at younger age, p<0.001) and dyslipidemia (14% vs. 3.4-9.1% at younger age, p-0.007), and significantly higher BMI percentiles, with 52% and 57% of patients in age groups 6-12 years and 13-18 years, respectively, having BMI values > 85^th^ percentile for age and sex, compared to patients under 6 years of age (p<0.001).

Table 4 shows the characteristics of patients diagnosed with primary and secondary HTN, indicating major demographic and clinical differences between these two subgroups. Patients with primary HTN were significantly older at diagnosis (median age 15.9 years vs. 9.4 years for secondary HTN, p<0.001), and had a higher prevalence of overweight and obesity (overweight in 19% and obesity in 69%, vs. 11% and 19%, respectively, for secondary HTN, p<0.001). The majority of primary HTN patients were Jewish (45% vs. 25% among secondary HTN, p<0.001), while the majority of secondary HTN patients were of Arab-Muslim ethnicity (51% vs. 28% for primary HTN, p<0.001). Family history of HTN was more prevalent in patients with primary HTN (26% vs. 7.1% for secondary HTN, p<0.001). Of the secondary HTN group, mortality rate during follow-up was 9.1%, vs. 1.3% in the primary HTN group (p-0.019). The rate of complications was significantly higher in patients with secondary HTN, in whom LVH was observed in 41%, vs. 2.5% in the primary HTN subgroup (p<0.001). Nevertheless, the prevalence of hypertensive retinopathy was comparable between primary and secondary HTN subgroups. During follow-up, 12% of secondary HTN patients suffered from hypertensive emergencies, compared to none in the primary HTN subgroup (p-0.001). Conversely, comorbidities were more prevalent in primary HTN patients.

**Table 4:** Characteristics of patients with primary vs. secondary HTN.

| Variable |  | Primary HTN<br>N=80 | Secondary HTN<br>N=198 | p-Value<sup>1</sup> |
| --- | --- | --- | --- | --- |
| Median Age (IQR) |  | 15.9 (13.9, 17.1) | 9.4 (2.7, 14.0) | <0.001 |
| Gender | Male n(%) | 58 (72%) | 119 (60%) | 0.052 |
|  | Female n(%) | 22 (28%) | 79 (40%) |  |
| Mortality rate n(%) |  | 1 (1.3%) | 18 (9.1%) | 0.019 |
| Consanguinity n(%) |  | 8 (10%) | 43 (22%) | 0.022 |
| Positive family history of HTN n(%) |  | 21 (26%) | 14 (7.1%) | <0.001 |
| Ethnicity | Jewish n(%) | 36 (45%) | 50 (25%) | <0.001 |
|  | Arab Muslims n(%) | 22 (28%) | 100 (51%) |  |
|  | Arab Christians n(%) | 6 (7.5%) | 7 (3.5%) |  |
|  | Druze n(%) | 0 (0%) | 14 (7.1%) |  |
|  | Other n(%) | 16 (20%) | 27 (14%) |  |
| BMI percentiles at diagnosis<sup>2</sup> | Obesity n(%) | 37 (69%) | 22 (19%) |  |
|  | Overweight n(%) | 10 (19%) | 13 (11%) |  |
|  | Normal weight n(%) | 7 (13%) | 69 (60%) |  |
|  | Underweight n(%) | 0 (0%) | 11 (9.6%) | <0.001 |
| Obesity diagnosis during follow-up n(%) |  | 59 (74%) | 29 (15%) | <0.001 |
| Prematurity <37 weeks' gestation n(%) |  | 5 (6.2%) | 21 (10.6%) | 0.3 |
| eGFR below 60 at diagnosis n(%) |  | 0 (0%) | 38 (38%) | <0.001 |
| Median eGFR at Dx (IQR) |  | 100 (91, 112) | 79 (29, 106) | <0.001 |
| Complications | LVH n(%) | 2 (2.5%) | 81 (41%) | <0.001 |
|  | Hypertensive retinopathy n(%) | 5 (6.2%) | 20 (10%) | 0.3 |
|  | Need for Dialysis during F/U n(%) | 0 (0%) | 84 (42%) | <0.001 |
|  | Proteinuria during F/U n(%) | 0 (0%) | 58 (46%) | <0.001 |
|  | Hypertensive emergency during F/U n(%) | 0 (0%) | 24 (12%) | 0.001 |
| Comorbidities | Diabetes Mellitus n(%) | 15 (19%) | 11 (5.6%) | <0.001 |
|  | Dyslipidemia n(%) | 18 (22%) | 25 (13%) | 0.039 |
|  | OSA n(%) | 5 (6.2%) | 7 (3.5%) | 0.3 |
|  | Hyperuricemia n(%) | 6 (15%) | 73 (42%) | 0.001 |
| Interventions | Vascular intervention<sup>3</sup> n(%) | 0 (0%) | 8 (4%) | 0.11 |
|  | Nephrectomy n(%) | 0 (0%) | 14 (7%) | 0.013 |

F/U= follow-up; LVH= left ventricular hypertrophy; OSA= obstructive sleep apnea
<sup>1</sup>Kruskal-Wallis rank sum test; Fisher's Exact Test for Count Data with simulated p-value (based on 1e+06 replicates); Pearson's Chi-squared test
<sup>2</sup> BMI percentiles: Underweight BMI<5<sup>th</sup> percentile, Normal weight BMI percentiles 5-85<sup>th</sup>, Overweight BMI percentiles 85-95<sup>th</sup>, Obesity BMI>95<sup>th</sup> percentile
<sup>3</sup> Renal artery embolization, dilatation, revascularization, surgical intervention

Comparison of demographic and clinical characteristics between hypertensive patients with kidney failure undergoing dialysis therapy, renal transplant recipients (RTR) and all other remaining patients in the cohort has shown no significant differences between subgroups in relation to age, gender, mortality rate, or BMI at diagnosis (table 5). In contrast, ethnicity differed significantly between subgroups, with dialysis patients and RTR originating more frequently from Arab-Muslim ancestry (70% and 63% respectively), compared to 35% Arab-Muslims in the rest of the cohort. Complication rate was also more prevalent in the dialysis and RTR groups, with LVH reported in 50-60% of patients, vs. 11% in the rest of the cohort (p<0.001). Dialysis patients and RTR had higher rates of hypertensive emergencies (18-19% vs 2.8% in the rest of the cohort, p<0.001). Dialysis patients and RTR underwent significantly more renal artery interventions and nephrectomies compared to the rest of the cohort.

**Table 5.** Characteristics of patients with ESRD compared to renal transplant recipients.

| Variable |  | ESRD<br>under<br>dialysis<br>N=90 | Renal<br>transplant<br>recipients<br>N=52 | Others<br>N=386 | p-Value<sup>1</sup> |
| --- | --- | --- | --- | --- | --- |
| Median Age (IQR) |  | 9.4 (3.0,<br>14.4) | 11.6 (2.6,<br>15.9) | 11.3 (5.1,<br>14.7) | 0.2 |
| Gender | Male n(%) | 33 (63%) | 50 (56%) | 252 (65%) | 0.2 |
|  | Female n(%) | 19 (37%) | 40 (44%) | 134 (35%) |  |
| Mortality rate n(%) |  | 13 (14%) | 32 (8.3%) | 5 (9.6%) | 0.2 |
| Ethnicity | Jewish n(%) | 12 (23%) | 15 (17%) | 138 (36%) | <0.001 |
|  | Muslim Arab<br>n(%) | 33 (63%) | 63 (70%) | 134 (35%) |  |
|  | Christian Arab<br>n(%) | 1 (1.9%) | 2 (2.2%) | 16 (4.1%) |  |
|  | Druze n(%) | 3 (5.8%) | 4 (4.4%) | 21 (5.4%) |  |
|  | Other n(%) | 3 (5.8%) | 6 (6.7%) | 77 (20%) |  |
| BMI<br>percentiles at<br>diagnosis<sup>2</sup> | Underweight<br>n(%) | 5 (15%) | 8 (18%) | 18 (9.4%) | 0.2 |
|  | Normal weight<br>n(%) | 19 (56%) | 23 (51%) | 80 (42%) |  |
|  | Overweight<br>n(%) | 3 (8.8%) | 3 (6.7%) | 29 (15%) |  |
|  | Obesity | 7 (21%) | 11 (24%) | 64 (34%) |  |
| Obesity diagnosis during follow-up n(%) |  | 14 (16%) | 101 (26%) | 10 (19%) | 0.008 |
| Complications | Left ventricular<br>hypertrophy<br>(LVH) n(%) | 26 (50%) | 52 (58%) | 41 (11%) | <0.001 |
|  | Hypertensive<br>retinopathy<br>n(%) | 3 (5.8%) | 9 (10%) | 21 (5.4%) | 0.3 |
|  | Hypertensive<br>emergency<br>n(%) | 10 (19%) | 16 (18%) | 11 (2.8%) | <0.001 |
| Comorbidities | Diabetes<br>Mellitus n(%) | 5 (9.6%) | 6 (6.7%) | 23 (6.0%) | 0.5 |
|  | Dyslipidemia<br>n(%) | 8 (15%) | 16 (18%) | 29 (7.5%) | 0.006 |
|  | OSA n(%) | 0 (0%) | 3 (3.3%) | 11 (2.8%) | 0.5 |
|  | Hyperuricemia<br>n(%) | 35 (69%) | 60 (67%) | 30 (11%) | <0.001 |
| Interventions | Vascular<br>intervention<sup>3</sup><br>n(%) | 3 (5.8%) | 4 (4.4%) | 4 (1.0%) | 0.012 |
|  | Nephrectomy<br>n(%) | 5 (9.6%) | 7 (7.8%) | 7 (1.8%) | 0.001 |

LVH= left ventricular hypertrophy; F/U= follow-up, OSA=obstructive sleep apnea
<sup>1</sup>Kruskal-Wallis rank sum test; Fisher's Exact Test for Count Data with simulated p-value (based on 1e+06 replicates); Pearson's Chi-squared test
<sup>2</sup> BMI percentiles: Underweight BMI<5<sup>th</sup> percentile, Normal weight BMI percentiles 5-85<sup>th</sup>, Overweight BMI percentiles 85-95<sup>th</sup>, Obesity BMI>95<sup>th</sup> percentile
<sup>3</sup> Renal artery embolization, dilatation, revascularization, surgical intervention

## Discussion

Over the past decades, the reported prevalence of pediatric HTN has gradually increased, posing a significant public health challenge, due to its long-term adverse effects. In this observational retrospective cohort study, we examined the EMR of 479 pediatric patients diagnosed with HTN at Rambam health care center, with 268 patients (56%) followed at the Pediatric Nephrology Institute.

The demographic and epidemiologic characters of this study population aligned with existing literature. Expectedly, the mean age of HTN at diagnosis was 9.6 years (±6.3 years), similar to 9.1 years reported by Kaelber *et. al*(2) in a large multi-center study. Similarly, 45% of our cohort had a BMI above the 85^th^ percentile for age and sex at diagnosis, comparable to previous reports of 54%(2) and 49%(9). Interestingly, abnormal BMI was more common in our cohort, compared to the 21% overweight and 33% obesity prevalence reported in children and adolescents of northern Israel in 2018(25). Notabley, males constituted 64% of our cohort, higher than the 47%-55% male prevalence reported in other studies (2,27).

Only 17% of patients in our cohirt were diagnosed with primary HTN, significantly lower than up- to 50% prevalence reported elswhere (28–32). Our rates of primary pediatric HTN aligh with a recent meta-analysis showing higher prevalence in primary care or school settings than in referral clinics(33). Nontheless, primary HTN prevalence in our tertiary care facility remains relatively low. These discrepancies may stem from underdiagnosis of HTN in community setting in our region, leaing to a higher referral rate for secondary HTN cases. Bureaucratic barriers may further limit referrals to hospital specialized HTN clinics, as evidenced by a recent study showing only 37.8% of community patients being diagnosed according to 2017 AAP guidelines, and just 5.5% referred to specialty HTN clinics(34). Community-based studies are needed to explore the lower rates of primary HTN referrals in our region.

Ethnic distribution and high consanguinity rates also contribute to the high rates of secondary HTN in our study population. Arab-Muslim ethnicity accounted for 42% of our cohort, higher than its 30.5% prevalence in northern-Israel(25). Consanguinity rates in this population reached 30%(35), contributing to the overall 16% consanguinity rate in the entire study cohort.

Autosomal recessive diseases are eight-times more prevalent in consanguineous populations of Israel, affecting 15% of non-Jewish Israeli infants(36). Many of the patients in our cohort suffer from underlying monogenic disorders linked to high consanguinity rates. Patients of Arab Muslim ethnicity were significantly younger, had lower eGFR, and exhibited higher complication rates, reflecting increased secondary HTN requiring close follow-up and strict blood pressure control.

Consistent with previous sudies, significant differences were observed between primary and secondary HTN subgroups (27,30,31,37,38). Our patients with primary HTN were older (median age 15.9 years vs. 9.4 for secondary HTN), comparable to previous reports by Rasala *et.* al(27) and others, predominantly Jewish, and had a higher prevalence of family history of HTN. Positive family history is more common in primary HTN(27,30,31,37,38), likely due to genetic predispositions and familial lifestyle factors. BMI over the 85^th^ percentile was found in 88% of primary HTN population, while obesity was found in 69%, aligning with the findings reported by Rasala *et.* al(27) and Gomes *et.* al(30). These distinctions aid in identifying at-risk patients and tailoring their work-up, management, and follow-up.

Drug-related HTN constituted 13% of our cohort, primarily secondary to corticosteroid use for conditions such as acute lymphoblastic leukemia (ALL), intractable seizures, and peri-extubation. Corticosteroids are well-known to cause HTN, affecting up-to 40% of patients treated with ACTH for west syndrome(39), and 45% during ALL induction therapy(40). Acute HTN, often pain or anxiety-related, accounted for additional 15% of cases, particulariy in the PICU setting.

Despite the 2017 AAP guidelines focusing on ambulatory HTN patients, our study highlights the challenges of diagnosing and managing HTN in hospitalized pediatric patients. Approximately 1% of general pediatric and 25% of PICU patients are estimated to have hospitalization-related hypertension, with significant underdiagnosis suspected(41). In our study, acute and drug-related HTN exhibited a 7.8% complication rate, including LVH in seven patients, Hypertensive emergencies in three, and both in one. These findings emphasize the importance of meticulous monitoring, diagnosis, and early intervention in hospitalized pediatric patients at risk for HTN.

Referral rates and diagnostic evaluations varied across the cohort. While the AAP recommends thorough investigation for HTN in patients under six years of age (Carroll *et. Al*)(34), community referral rates remain low, at just 7.9% for this age group. In our hospital-based study, 52% of patients under six were referred to the Pediatric Nephrology Institute, with 57.8% undergoing a thorough evaluation for secondary HTN. Across all age groups, 53.4% of patients received such evaluations. Given the high prevalence of genetic diseases in our population due to consanguinity, comprehensive investigations are warranted for all age groups, and should not be restricted to younger patients.

These results emphasize the importance of specialized follow-up and treatment for secondary HTN patients to prevent complications.

ABPM was more frequently utilized in the nephrology subgroup. ABPM plays a critical role in monitoring EBP, as it is better correlates with EOD(22). Our results demonstrate that pediatric HTN poses a significant risk for EOD, particularly in the nephrology subgroup and among secondary HTN patients. LVH, a preventable risk factor for future cardiac disease(37) was significantly more prevalent in these groups. Moreover, hypertensive emergencies, although rare in children, occurred in 12% of secondary HTN patients, but none with primary HTN, mirroring findings from Gomes *et.* al(30). These results emphasize the importance of meticulous follow-up and treatment for secondary HTN patients in specilized pediatric nephrology clinics, to prevent complications and EOD.

Pharmacological management of pediatric HTN remains challenging, due to limited evidence(42), and lack of FDA approved anti-hypertensive medications for young children(43). Current guidelines do not specify preferred drug class, leaving treatment decisions to individual practitioners. RAAS inhibitors are most commonly prescribed in pediatric HTN (5,44), with Kaelber *et. al*(2) reporting 35% useage compared to 18% for CCB. However, in our cohort, CCBs were used in nearly 80% of patients, while RAAS inhibitors were prescribed in only 28%. The high prevalence of vascular HTN , CKD, RRT and RTR in our cohort likely explains our widespread use of CCBs. Of note, however, incomplete documentation of treatment in non-nephrology units may introduce selection bias in our results.

Second, retrospective EMR-based inclusion criteria may lead to underdiagnosis of HTN. Our study has several limitations. First, our data is derived from a tertiary referral center and may be subject to referral bias, limiting its applicability to community setting. In addition, retrospective EMR-based inclusion criteria may lead to underdiagnosis of HTN.

In conclusion, this study highlights the unique demographic, cultural, and environmental factors influencing the prevalence, etiology, severity, and management of pediatric HTN in a large tertiary referral center in northern Israel. Our findings emphasize the importance of adherence to guidelines and the benefits of specialized care at a pediatric HTN clinic. While standardized guidelines are essential, flexibility is necessary to address the specific needs of diverse populations under care.

## Disclosure

All authors have contributed equally to the work.

The authors declare no conflicts of interest.

Ethical board approval was given by Rambam Health Care Campus Helsinki committee number 0127-21

All data generated or analysed during this study are included in this published article, more data that support the findings of this study are available from the corresponding author upon reasonable request.

## Data Availability

All data generated or analyzed during this study are included in this published article, more data that support the findings of this study are available from the corresponding author upon reasonable request.

## Statements and Declarations

The authors declare that no funds, grants, or other support were received during the preparation of this manuscript.

The authors have no relevant financial or non-financial interests to disclose.

All authors wrote the main manuscript text, L.W. prepared the tables and figures, all authors reviewed the manuscript.

This study was performed in line with the principles of the Declaration of Helsinki. Approval was granted by the Ethics Committee of Rambam Health Care Campus, nomber of approval: RMB-0127-21.

According to the Helsinki committee as the data collection from the patients’ files using a data extraction platform), we were authorised to proceed without the need for informed concent.

The data that support the findings of this study are not openly available due to reasons of sensitivity and are available from the corresponding author upon reasonable request. Data are located in controlled access data storage at Rambam Health Care Center

